# Mortality Doubled After Stressful Football Match

**DOI:** 10.1101/2022.09.07.22278977

**Authors:** Roland Rau, Marcus Ebeling, Bernhard Köppen

**Author notes:** Corresponding Author: University of Rostock, Chair of Demography, Ulmenstr. 69, 18057 Rostock, Germany.

## Abstract

**Background:** Strong emotional stress can lead to death. Even less serious events such as watching stressful football matches are proposed as triggers for mortality. The results are inconclusive, though, probably due to a focus on national teams. Instead, this study analyzes daily mortality after a stressful match of a club team. It also accounts for the role of temperature, a major confounder for daily mortality.

**Methods:** Daily deaths from 01 January 1999 until 10 July 2020 were used to predict mortality separately for women and men on the day of the match (11 July) and the subsequent days. The time series regression model assumed a negative binomial distribution for the death counts, accounted for potential autocorrelation and included temperature as a regressor. 95% prediction intervals were estimated via bootstrapping.

**Results:** Two days after the match, mortality was twice as high as expected for men. In contrast to the analysis for women, temperature can be ruled out as a contributing factor.

**Conclusion:** Watching stressful football matches can be a matter of life and death for susceptible persons. Making people aware of it could be a cost-efficient way to lower the burden of dying from circulatory diseases.

## Introduction

Sudden emotional stress can lead to cardiovascular diseases and potentially to death. Measuring the impact of stress on mortality is challenging since the consequences of a stress inducing event are typically closely related to the individual’s perception of stress [1].

Testing the lethality of stress on the population level requires a triggering event affecting many people at the same time. Earthquakes are an example [2]. But even apparently less grave incidents—such as watching a football match—are proposed. The results from past research are inconclusive, though [e.g., 3–9]. Most of these studies focus on games at World Cups or European Championships. We argue that this is partially misguided: While more people are watching the games of a national team, it is suggested that identification with a football club — particularly if there is a strong local tie to the club — can be more important to individuals [10]. A dramatic course in a landmark game of the supported club would therefore represent a more suitable trigger mechanism. That is why it should not be surprising that one of the studies showing a strong effect on mortality is based on club football instead of an international competition [11]. Similar to other studies, however, it fails to control for other potential confounders, most notably temperature, which can have a major impact on daily mortality [12].

We analyzed daily mortality in Nuremberg, a city of about 500,000 inhabitants in the Northern part of Bavaria, Germany, in the aftermath of a stressful football match, taking temperature into account: The city is home to Nuremberg Football Club (“FCN”). The team has the second most national titles in Germany and a large and loyal regional fan base. In the evening of 11 July 2020, the second and final leg of the playoffs were played between FCN and FC Ingolstadt to determine promotion and relegation between the second and third division. Being relegated to the third division would have been a disaster in the eyes of the supporters of FCN. The game had a dramatic finish: FCN scored the decisive goal to stay in the second division about 30 seconds after the officially allotted extra time of already five minutes. The tension of the game was so tangible that a renowned journalist published a book entitled “Football as a near-death experience” afterwards [13]. We argue that the emotional stress and unexpected relief due to the last-second goal represents an excellent setting to analyze the effects of emotional stress on daily mortality.

## Methods

We obtained daily death counts for the time period 01 Jan 1999 through 31 July 2020 from the statistical office of Nuremberg [14]. During that time span of 7,883 days, 121,176 individuals passed away. 53% were women (64,275) and 47% were men (56,901 men). Data for the mean daily temperature for Nuremberg were obtained from the “Deutscher Wetterdienst” [15, station id for Nuremberg: 03668] for the corresponding time period.

Daily deaths were fitted with R [16] and the extension package `tscount` [17] until the day before the match (01/01/1999–10/07/2020) by using a log-link and assuming a negative binomial distribution to allow for overdispersion in the count data. Potential dependence across observations (e.g., autocorrelation) was accounted for by regressing on past observations and past conditional means. To avoid selecting an arbitrary model, we conducted an extensive grid search for both components starting at zero (no dependence structure) to three. The impact of temperature was measured a) directly on the given day, b) on one through five previous days, or c) as the change in temperature during the past one through five days. This led to eleven estimated models for each given dependence structure. Each fitted model was then used to predict daily deaths for the subsequent seven days, starting on the day of the match (11/07/2020). 95% prediction intervals were obtained using a parametric bootstrap procedure with 500 replications, which is sufficient for stable results.[17].

The ethics committee of the University of Rostock approved the study (A 2022-0141).

## Results

The highest number of male deaths during the predicted time interval was 15. It was observed on 13 July 2020. Such a lag period of up to two days between trigger and death matches also the experience of the Athens earthquake [2]. The left panel of Fig. 1 shows how exceptional 15 deaths are for men during that time of the year (13th July ± 14 days in 1999–2020): There was only a single day ever with more deaths during these 638 days.

**Figure 1:**
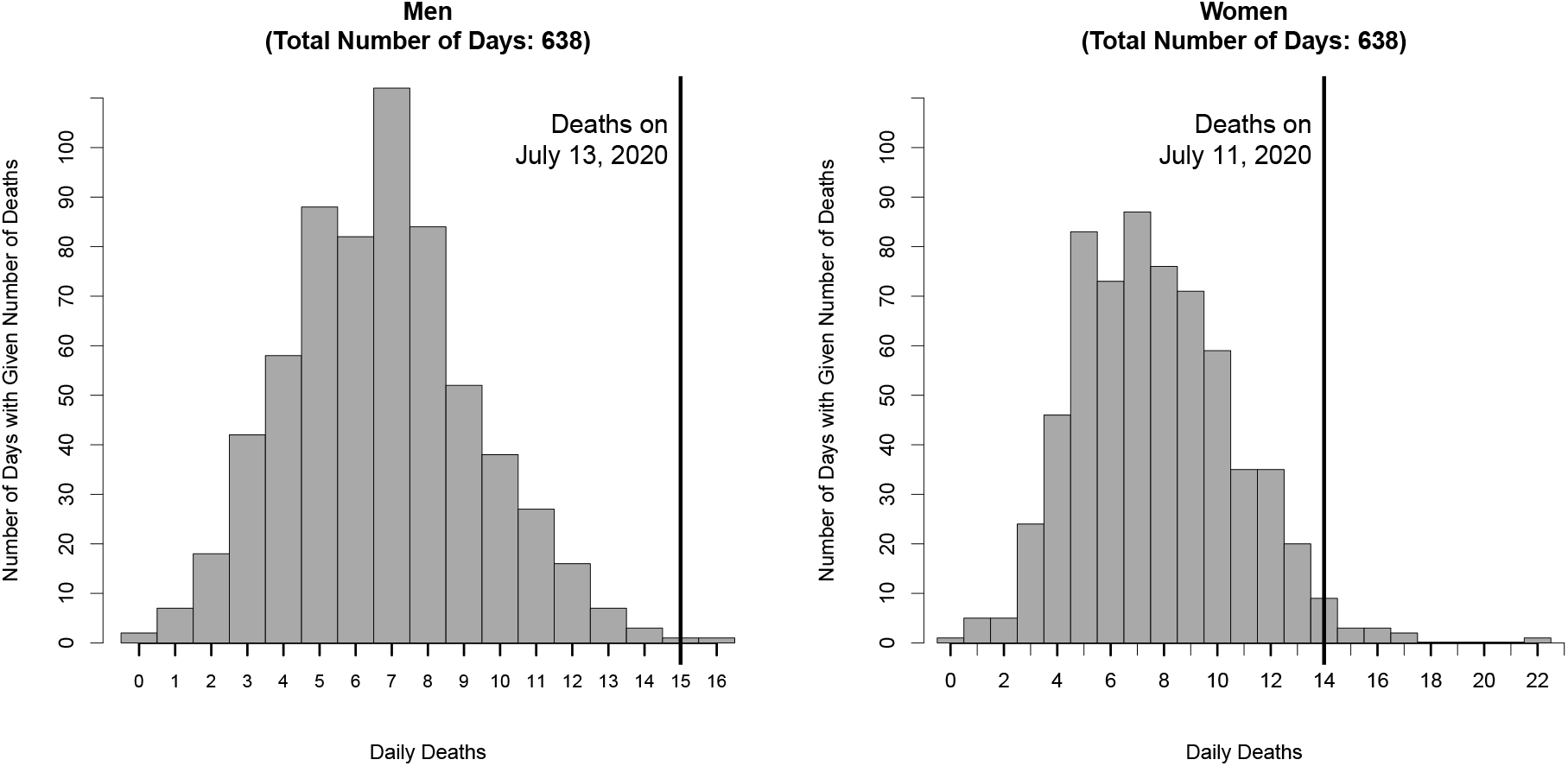
Histogram of daily deaths in Nuremberg for men on July 13 ± 14 days (left panel) and for women on July 11 ± 14 days (right panel) during 1999–2020. Vertical reference lines have been added for the respective day with the highest number of deaths within a week following July 10, 2020.

The highest number of deaths for women was recorded on the day of the match (14 deaths). While still being high, it is not as exceptional as shown in the right panel of Fig. 1. Since the match finished at about 8pm, we believe it is unlikely that the elevated number of female deaths can be attributed to the football match, but rather to temperature: Mean daily temperature in Nuremberg rose from 14.8C to 22.4C in the days leading up to the match. Such a rise in temperature by 7.6C within three days in Nuremberg during that time of the year is extremely rare (98th percentile). Whether it may have induced stress on the circulatory system leading to additional deaths is shown in Fig 2.

**Figure 2:**
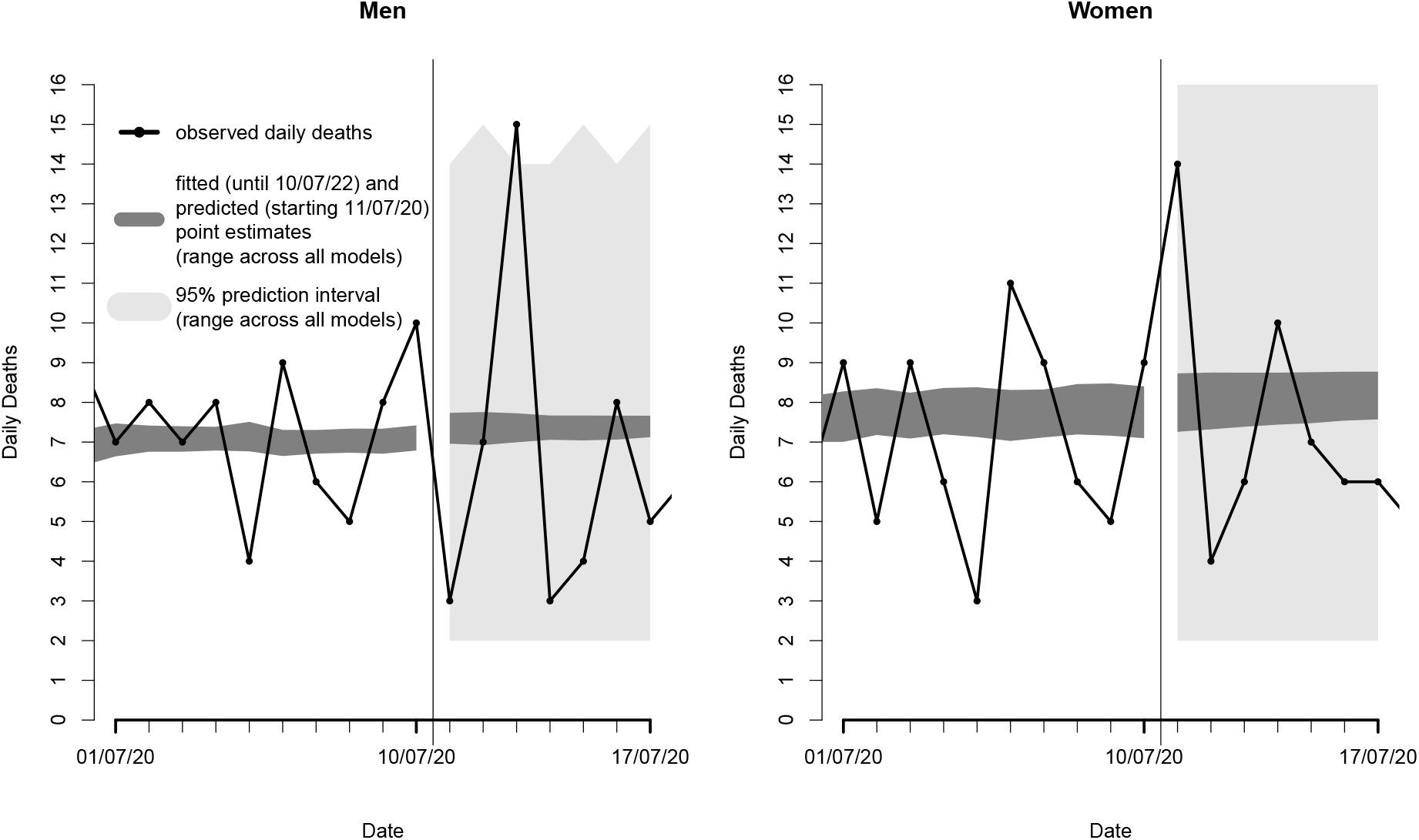
Observed, fitted (until 10/07/2020) and predicted (starting on 11/07/2020) daily deaths in Nuremberg for men (left panel) and women (right panel). The dark grey area depicts for each day the range of fitted (starting on 01/01/1999) and predicted point estimates across all models. The light grey area illustrates the corresponding range of maximum and minimum values from 95% prediction intervals across all models for each day. A vertical reference line has been added to separate fitting from prediction.

Observed deaths for men (left panel) and women (right panel) are depicted by a solid black line with black dots for each day. The dark grey area left of the vertical reference line denotes the range of point estimates across all models. The dark grey area to the right of the vertical reference line illustrates the range of predicted deaths for seven days, starting on the day of the match. The bright grey areas show the minimum and maximum values of the 95% prediction intervals across all estimated models. The observed number of 15 male deaths on 13th July is about twice as high as the predicted value of around seven. The excess mortality among women is almost as high (14 observed deaths vs. 7–8 expected deaths). However, the light grey area reaches 16 deaths on each day, indicating that the 95% prediction interval of at least one model covered the observed female deaths (68 model predictions included 14 or more deaths). Hence, we can not exclude the possibility that temperature may have played a crucial role for female excess mortality.

The situation is different for men: Not a single 95% prediction interval of any of the 176 estimated models covers 15 male deaths on 13th July 2020. We did not discover a trough in deaths in the days after the peak (“harvesting effect”), suggesting that the excess deaths were additional deaths and did not originate from frail persons who would have probably died within the next few days.

## Discussion

Watching stressful football matches can be a matter of life and death: We have shown that male mortality doubled in Nuremberg, Germany, two days after a stressful game. In contrast to the results for women, we can exclude temperature to have played a crucial role for the high number of male deaths. Additional information on cause of death—particularly due to circulatory diseases—would have been helpful but was not available. Nonetheless, we are confident that the football match triggered some of the excess male deaths: Not a single model included 15 deaths in their 95% prediction intervals despite a) checking 11 potential effects of temperature, b) 16 different dependence structures to account for autocorrelation, and c) using a negative binomial distribution instead of the more restrictive but canonical choice of a Poisson distribution for death counts.

We further tested the robustness of our predictions by conducting additional analyses with classic ARIMA time-series analysis [18], using function `auto.arima()` for best model selection, and Facebook’s automated “prophet” forecasting package.[19]. None of these analyses would have led to different conclusions.

Although pertained to club football, we want to put our results into a bigger context: An apparently unimportant event, inducing a high dose of emotional stress, can be enough to lead to death. It is likely that the deceased individuals have been more vulnerable than others due to some preexisting disposition or the presence of mortality risk factors. Most of these risk factors, such as obesity, exert continued damage to the body. Our results suggest that trigger events could play a crucial role in modulating these health deficits, a mechanism that might be most relevant for cardiovascular conditions. While a full understanding of the mechanisms requires also an analysis of patient characteristics and other outcomes such as hospital admissions and causes of death, making people aware of the potential dangers could be already a cost-efficient way to lower the burden of dying from circulatory diseases.

## Supporting information

Strobe Statement

## Data Availability

The data can be requested from the Stadt Nürnberg. Amt für Stadtforschung und Statistik, Unschlittplatz 7a, 90403 Nürnberg, Germany. To obtain the identical data, please request the tägliche Nürnberger Sterbefallzahlen in den Jahren 1999 bis 2020.

**WHAT IS ALREADY KNOWN ON THIS TOPIC**

Past studies reported increased mortality levels in the days after a stress inducing event, such as an earthquake. Similar observations have been made in the aftermath of dramatic sports competitions, such as football matches. The findings illustrated the lethal potential of emotional stress.

**WHAT THIS STUDY ADDS**

Stress induced by a dramatic course of a sports event can lead to exceptionally high mortality in the days thereafter. Our findings suggest that the level of stress is likely modulated by the bonds between the fanbase and “their” team, particularly on the regional level. Our finding emphasizes the potential of apparently unimportant events for triggering short-term increases in mortality and excess deaths. It remains an open question to what extent such situations are also a challenge for local health care infrastructure.

## Data availability statement

The authors are not allowed to share the data. The data can be requested from the “Stadt Nürnberg. Amt für Stadtforschung und Statistik”, Unschlittplatz 7a, 90403 Nü rnberg, Germany. To obtain the identical data, please request the “tägliche Nürnberger Sterbefallzahlen in den Jahren 1999 bis 2020”.

